# The appropriateness of empirical antibiotic therapy in the management of symptomatic urinary tract infection patients-A cross sectional study in Nairobi County, Kenya

**DOI:** 10.1101/2024.06.19.24309164

**Authors:** Hellen A. Onyango, Derek J Sloan, Katherine Keenan, Mike Kesby, Caroline Ngugi, Humphrey Gitonga, Robert Hammond

**Affiliations:** School of Medicine, University of St Andrews, Scotland, United Kingdom; School of Geography and Sustainable Development, University of St Andrews, Scotland, United Kingdom; College of Health Sciences, Jomo Kenyatta University of Agriculture and Technology, Nairobi, Kenya; Centre for Microbiology Research, Kenya Medical Research Institute, Nairobi, Kenya

**Author notes:** Corresponding author: Hellen A. Onyango.

## Abstract

**Background:** In low- and- middle income countries, symptomatic urinary tract infection (UTI) patients are often prescribed antibiotics without microbiological confirmation. UTIs caused by antibiotic resistant bacteria are increasingly common and this heightens the risk of empirical treatment failure. This study evaluates the appropriateness of empirical antibiotic therapy to symptomatic UTI patients.

**Methods:** A hospital-based, cross-sectional study was conducted in Nairobi County, Kenya among symptomatic adult and child patients. UTI was defined as a monoculture growth with colony counts of ≥10^4^. cfu/ml. Antimicrobial susceptibility testing (AST) was performed by the Kirby-Bauer disc diffusion method. Empirical therapy was considered appropriate if the pathogen isolated was susceptible to the prescribed antibiotic and inappropriate if pathogen was resistant to prescribed antibiotic.

**Results:** A total of 552 participants were enrolled with a median age of 29 years (IQR:24-36). The majority were female; 398 (72%). Of the 552, 274 (50%) received empirical antibiotic therapy, 95/274 (35%) were confirmed to have UTI by culture. The antibiotics most frequently prescribed were fluoroquinolones [ciprofloxacin in 80 (30%) and levofloxacin 43 (16%)], amoxicillin-clavulanic-acid in 48 (18%), and nitrofurantoin in 32 (12%). Amongst the 95 patients with bacteriological confirmation of UTI, 50 (53%) received appropriate empirical antibiotic therapy, whilst for 38 (40%) participants, the therapy was inappropriate. Appropriateness of empirical therapy to 7 (7%) patients could not be determined as the antibiotics prescribed were not in the AST panel.

**Conclusion:** The complexity of appropriate empirical treatment for UTIs is compounded by high levels of resistance in UTI pathogens. AMR surveillance strategies that could help in designing appropriate empirical regimens in resource constrained settings should be adopted for optimal empiric therapy.

## 1. Introduction

Antimicrobial resistance (AMR) is the ability of microorganisms to circumvent the toxic action of antimicrobial substances that otherwise would kill or inhibit them.^1^ The prevalence of resistance in common disease-causing bacteria has increased globally, both in healthcare and community settings.^2^ Consequently, the WHO has now listed AMR as an emerging public health threat believed to account for over 700,000 deaths per year.^3^ The burden of AMR is estimated to be highest in low-and-middle income countries (LMICs), particularly in Africa,^4^ where morbidity and mortality from infectious diseases is high, and health facilities less well-resourced than those in high income regions.^5,6^ Large regional, interdisciplinary studies, including the Holistic Approach to Unravel Antimicrobial resistance in East Africa (HATUA) project which was run across Kenya, Uganda, and Tanzania, have reported multiple drivers of AMR. Relevant factors ranged from inappropriate antibiotic prescriptions, to widespread non-prescription-based dispensing of antimicrobials for self-medication, antibiotic use in animals, environmental factors such as sanitation, as well as social-economic and structural drivers including the cost of seeking healthcare.^7–9^ In hospital settings, factors such as inadequate diagnostic capabilities, poor antibiotic stewardship practices, poor adherence to treatment guidelines and lack of AMR surveillance have been associated with resistance.^6,10^

Urinary tract Infections (UTIs) are among the most common community-acquired bacterial infections, and are the second most frequent clinical indication for antibiotic use.^11^ after respiratory infections.^12^ Patients with suspected UTI are often initiated on antibiotic treatment before culture results are available. However, in some cases, approximately 40% of the bacteria that cause UTI are resistant to the antimicrobials prescribed.^13^ In the recent past, the prevalence of multi-drug resistant (MDR) bacteria associated with UTI has increased,^14^ making selection of therapy for community acquired UTI complex. Guidelines for uncomplicated UTI treatment recommend customization of therapy based on local practice, circulating resistant organisms, drug availability and price.^15^ In Nairobi County, Kenya where this study was undertaken, nitrofurantoin 100mg, amoxicillin-clavulanic-acid 625mg, and amikacin 15-30mg/kg are recommended empirical antimicrobial therapy for community acquired UTIs.^16^ For recurrent infections, the guideline recommends that empirical therapy be guided by previous culture results pending urine culture and sensitivity results. Once available, therapy is tailored to prescribe the most narrow spectrum efficacious antibiotic wherever possible.^16^

The selection of empirical therapy for UTI management is dependent on knowledge of circulating pathogens and their antimicrobial resistance patterns.^17^ Of concern, therefore, is the lack of susceptibility data for community acquired UTI’s in many LMICs, including Kenya. This is mostly due to challenges with culture and susceptibility testing, some of which include infrastructural constraints, limited funding, prolonged turnaround times (TAT), and lack of skilled personnel.^4,18^ There is relatively limited information on the appropriateness of empirical antibiotic therapy in the management of community acquired UTI’s in LMICs. This study seeks to address the paucity of microbiological information on management of microbiologically confirmed UTI in symptomatic patients and evaluates the appropriateness of empirical UTI treatment based on culture and susceptibility results.

## 2. Materials and Methods

### 2.1. Study design

A hospital-based cross-sectional study design was employed to recruit adult and child patients with UTI-like symptoms between July 2022 to April 2023.

### 2.2 Study setting

The participants were recruited from Mama Lucy Kibaki Hospital (MLKH) and Mbagathi County Hospital (MCH) located within Nairobi County, as shown in **Figure 1**. The Kenyan healthcare system is structured in a hierarchical manner consisting of six levels I-VI in ascending order. MLKH and MCH are Level - V public referral hospitals. MCH and MLKH serve a large catchment area comprising both the middle and low socio-economic groups. The two hospitals were selected as there was limited information on the resistance profiles of circulating uropathogens and UTI patients are often treated empirically without culture confirmation.

**Figure 1:**
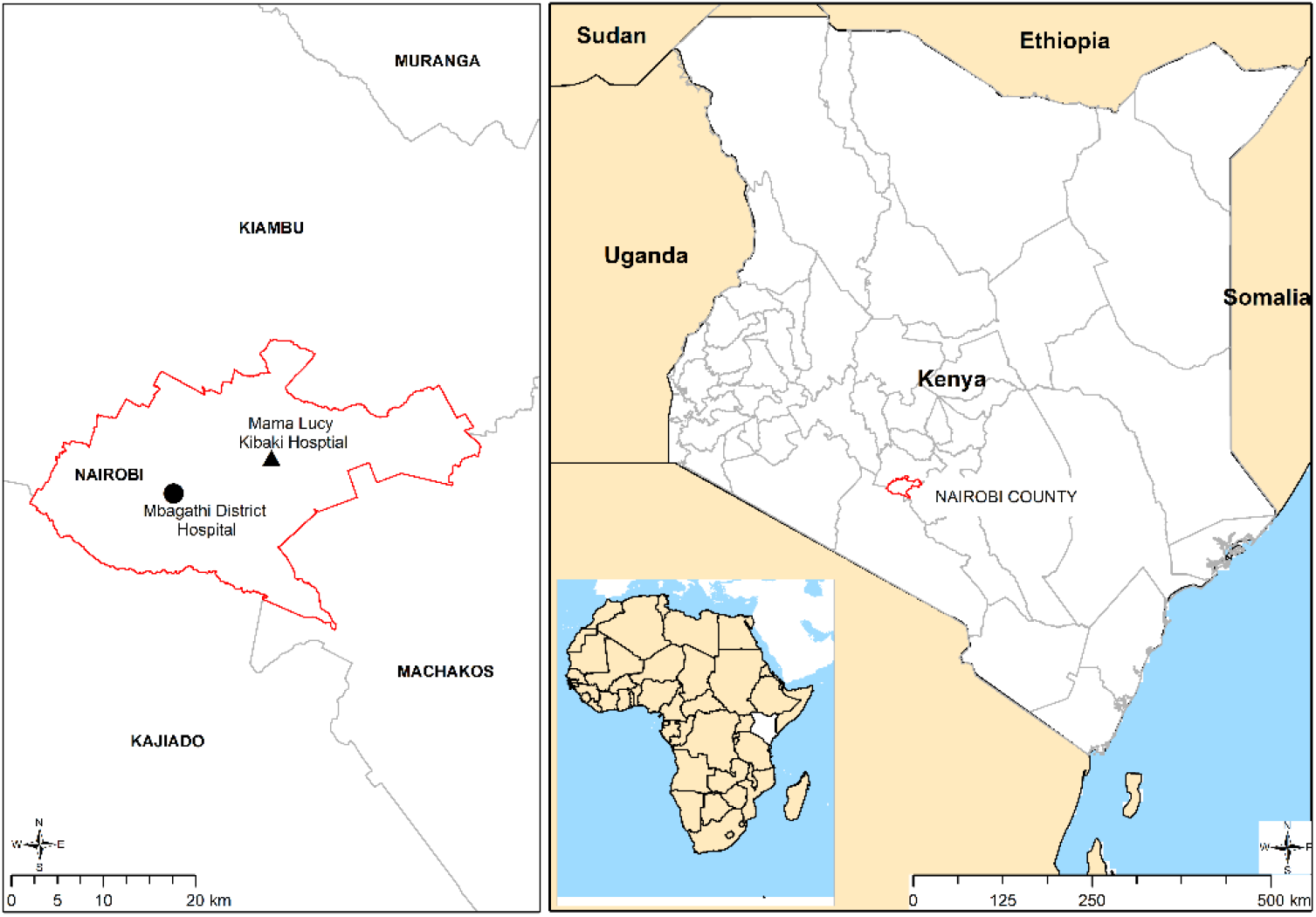
Location of study sites, MLKH and MCH within Nairobi County, Kenya.

### 2.3 Participant recruitment and sample collection

A resident clinician identified adult (≥18 years) and child (5-17 years) outpatients presenting with one or more symptoms suggestive of UTI or for other causes that made the clinician to believe they might also have a UTI. The symptoms included lower abdominal pain, dysuria, strong persistent urge to urinate, haematuria, frequent micturition and/or unexplained fever (≥38°C), persistent irritability, and suprapubic pain/tenderness to palpation in children. In addition to meeting the criteria of a presumptive UTI case, the participants had to meet the following criteria: report living within a 50 km radius from the hospital facility, have a mobile telephone number, and be able to speak/understand/write either English or Kiswahili. The study objectives were explained and patients willing to participate were taken through informed consent document (ICD) in their preferred language. Consent was obtained from adult patients (≥18 years). Assent and consent were obtained for participants aged 13-17 years. Parents/ guardians of participants aged < 13 years consented on their behalf. Consenting participants signed and dated the consent forms. Participants/guardians who were not able to sign marked the consent with a thumb print. Consenting participants were issued with a unique identifiable number which linked their bar-coded consent form, demographic data questionnaire, and urine sample collection container. Self-collected midstream urine on a 20 mL sterile plain screw-capped universal bottles was obtained from each patient after guidance on the collection procedure. Parents/guardians were guided on how to collect midstream urine from their children. The samples were stored in a cool box (4°C) and transported to the Kenya Medical Research Institute (KEMRI) laboratory for processing within 2 hours.

### 2.4 Data collection

A questionnaire was used to collect self-reported demographic information (age, gender), and previous antimicrobial use. Data regarding empirical antibiotic treatment was obtained from prescriptions administered to the patients during the hospital visit. All data were captured electronically into an epicollect database (https://five.epicollect.net is a free open-source datacollection tool) and later linked to the laboratory urine culture results.

### 2.5 Microbiological tests

#### 2.5.1 Urine culture

Using a standard sterile loop, an aliquot (10µl) of urine was plated directly on Cystine lactose electrolyte deficient agar (CLED), blood agar (BA), and MacConkey agar (Oxoid, Basingstoke, UK), and incubated aerobically at 35-37°C for 24 hours. After an overnight incubation, quantification of colony forming units (cfu’s) was done by counting the number of colonies on a plate and multiplying by the dilution factor, as previously described by Miles and Misra.^19^ Pure bacterial growth yielding colony counts of ≥10,000 (10^4^) cfu/ml was interpreted as a confirmed UTI case. A mixed culture (with more than one colony type) or growth of <10,000 (10^4^) cfu/ml were non-confirmatory for UTI.

The organisms were identified to the species level using colonial morphological characteristics on CLED, BA, MacConkey agar (Oxoid, Basingstoke, UK), Gram-stain (Sigma-Aldrich, USA) and standard biochemical tests. Sulfide Indole Motility test, Methyl Red, Oxidase, Urease, Triple Sugar Iron, and citrate utilization was used to identify Gram negative organisms.^20^ Coagulase, Catalase, and haemolytic patterns on BA was used to confirm the presence of Gram-positive bacteria. Where necessary, the analytical profile index (API 20E) test was used to confirm the identity of strains following the manufacturer’s guidelines (BioMerieux, Charbonnieres, LesBains, France).

#### 2.5.2 Antimicrobial susceptibility test

Antimicrobial susceptibility testing (AST) was performed according to the Kirby Bauer disc diffusion method.^21^ The panel of antibiotic discs (Oxoid, Basingstoke, UK) tested included first line; amoxicillin-clavulanic-acid (20/10 μg), nitrofurantoin (300 μg), sulfamethoxazole trimethoprim (23.75/1.25 μg), and second-line; ciprofloxacin (5 μg) antibiotics used in the treatment of UTI as per local practice.^16^ Other antibiotics included in the panel were ceftazidime (30 μg), ceftriaxone (30 μg), cefepime (30 μg), cefoxitin (30 μg), gentamycin (10 μg), cefuroxime (30 μg), erythromycin (15 μg), and linezolid (30 μg). Susceptibility or resistance to the tested antibiotics were determined using the zone diameter interpretative criteria (breakpoints) according to the CLSI guidelines.^22^ Isolates that showed intermediate resistance to a given antibiotic were interpreted as resistant to that antibiotic. *E. coli* (ATCC-25922) and *S. aureus* (ATCC-25923) were used as quality control organisms to validate antibiotic discs potency and quality of the test media.

### 2.6 Evaluation of the appropriateness of empirical treatment

Appropriateness of empiric treatment was assessed by evaluating the treatment prescribed during the initial hospital visit with the subsequent laboratory urine culture and susceptibility results. The hospital visit during which the patient was recruited, and urine sample obtained, was defined as the initial visit. Empirical treatment was taken as any antibiotic treatment prescribed to the patient during the initial visit prior to urine culture results. Appropriateness was assessed on an individual patient basis for those patients whose urine specimen yielded significant bacterial growth for UTI (≥10^4^ cfu/ml). Appropriate empirical antibiotic therapy (AEAT) was considered if a UTI was confirmed on urine culture and the antibiotics prescribed were effective in inhibiting growth of the isolated pathogen *in vitro.*^23^ Inappropriate empirical antibiotic therapy (IEAT) was defined as UTI confirmed on laboratory culture, but with an isolated pathogen which was resistant to the antibiotic prescribed *in vitro.*^23^ AEAT was expressed as the percentage of patients with a culture positive urine specimen, and isolated pathogen tested as sensitive to the antibiotic prescribed. Conversely, IAET was expressed as the percentage of patients with a culture positive urine specimen who had an empiric prescription for which the isolated pathogen was tested as resistant.

### 2.7 Statistical analysis

Data were downloaded from epicollect into Microsoft Excel (Microsoft Corp, Redmond, Washington, USA) and were analysed using STATA 16 (StataCorp. 2019. Stata 183 Statistical Software: Release 16. College Station, TX: StataCorp LLC). The questionnaire data were linked to urinalysis, empirical prescription and AST data using anonymous patient identifiers. Baseline characteristics of the study population were reported as median (interquartile range [IQR]) for age, or as counts and percentages for categorical data. Differences between categorical variables were compared using the χ2 test or Fishers exact test where applicable. Statistical significance was considered at probability value of < 0.05.

### 2.8 Ethical approval

This study received approval from University of St. Andrews Teaching and Research Ethics Committee, United Kingdom [Approval no. MD15749]; Jomo Kenyatta University of Agriculture and Technology Institutional Ethics Review Board, Kenya [Approval no. JKU/IERC/02316/0166], National Commission for Science Technology and Innovation, Kenya [Approval no. P/21/12520]. Nairobi Metropolitan services, MLKH and MDH provided approvals for access to the study sites. Informed consent was obtained from each participant included in the study.

## 3. Results

### 3.1. Characteristics of study participants

Participants characteristics are shown in **Table 1**. Five hundred and fifty-two were enrolled. The majority were adult outpatients 494 (89.4%), and females accounted for 398 (72%). The most frequent age bracket was 21-30 with a median age of 29 years (IQR:24-36). Among the 552 enrolled patients, 236 (43%) had taken medication 2 weeks prior to enrolment, 168 (71%) of these had taken antibiotics, while 68 (29%) had taken medications other than antibiotics.

**Table 1:**
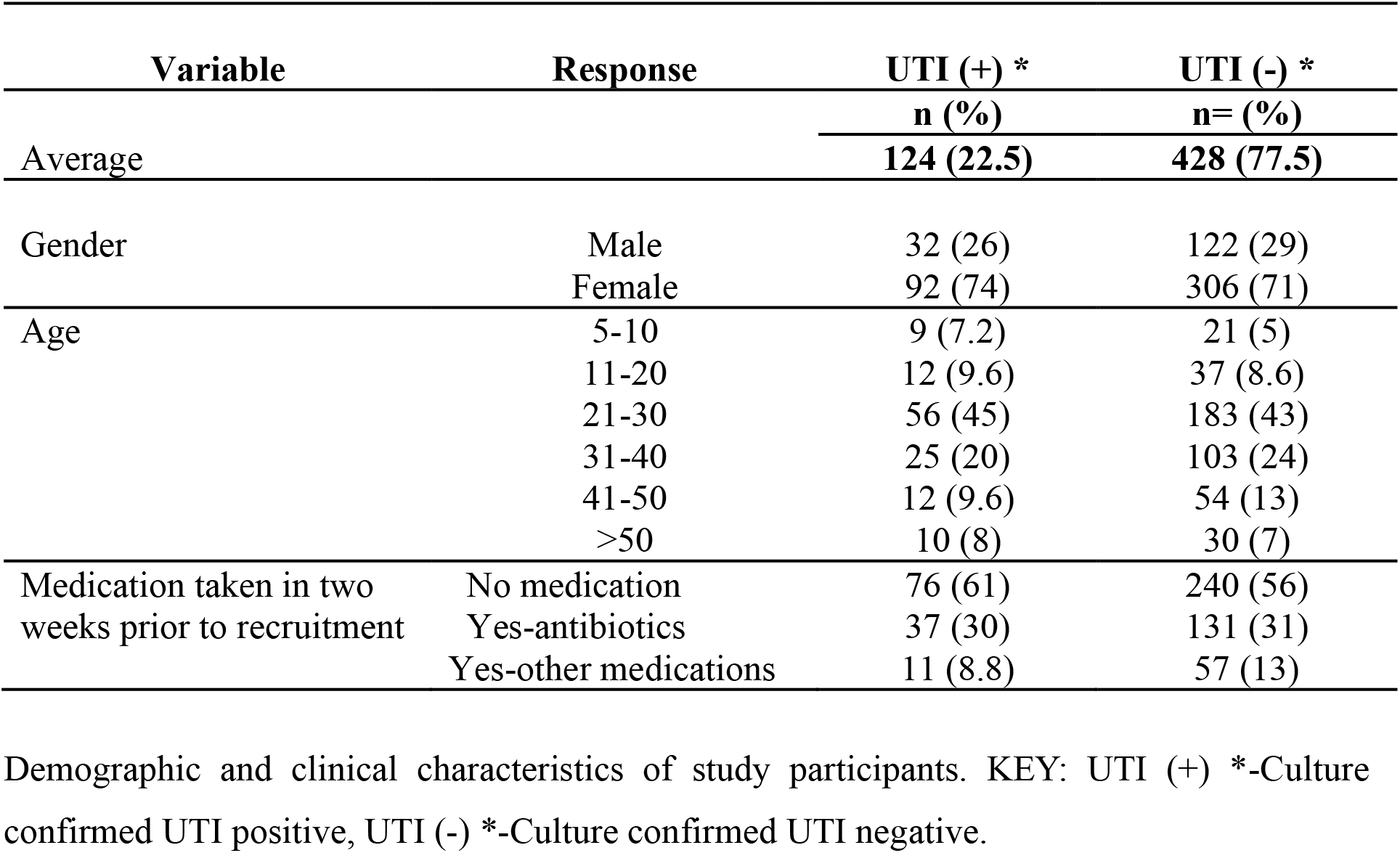
Basic demographic characteristics of participants.

### 3.2 Proportion of microbiologically confirmed UTI

The overall proportion of culture confirmed UTI among the studied population was 22.5%, (124/552), being significantly higher in females than males, **Table 1**. Of these, 274 (49.6%) received empirically prescribed antibiotic treatment, 242 (43.8%) did not receive any antibiotic treatment, while for 36 (6.5%), it was not known whether they received an antibiotic or not (participants could not be reached by phone or failed to come back to the hospital for the laboratory results). Amongst the 274 that received empirical antibiotic therapy, urine culture confirmed UTI in 95 (35%). Of the 242 that didn’t receive therapy, 27 (11.1%) had UTI confirmed. Among those whose therapy status was not known, 2 (5.5%) had confirmed UTI. There was a significant difference in UTI detection between those who received empirical therapy and those who did not, p-value of <0.05.

### 3.3 Microbiological characteristics

A total of 124 bacterial isolates were characterised from the 552 urine samples analysed, 97 (78%) of which were Gram-negative. The predominant uropathogen was *Escherichia coli* 64 (52%); followed by *Klebsiella* spp. 21 (17%); *Staphylococcus aureus*, 14 (11.3%); coagulase negative Staphylococcus (CoNS), 7 (5.6%); *Enterococcus faecalis*, 6 (4.8%), *Proteus* spp. 7 (5.6%); and *Acinetobacter baumanni*, 1 (0.8%). *Pseudomonas aeruginosa* 2 (1.6), and *Citrobacter koseri* 2 (1.6%).

### 3.4 Antimicrobial resistance patterns

Antimicrobial resistance profiles of the 124 isolated UTI pathogens are shown in **Table 2**. For Gram negative organisms, resistance towards common UTI treatments -β-lactams, fluoroquinolones, and aminoglycosides (BFQA) ranged from 24% to 57%. Within the bacterial groups, *E. coli*, the predominant uropathogen, showed high resistance to sulfamethoxazole/trimethoprim at 77%, ciprofloxacin at 61%, amoxicillin-clavulanic acid at 47%, and ceftriaxone at 52%, while nitrofurantoin was the most effective agent for *E. coli*. The overall resistance of Gram-positive bacteria was 52% for sulfamethoxazole trimethoprim, 67% for ciprofloxacin, and 26% for amoxicillin-clavulanic-acid. Nitrofurantoin and linezolid were the most effective agents against Gram-positive isolates.

**Table 2:**
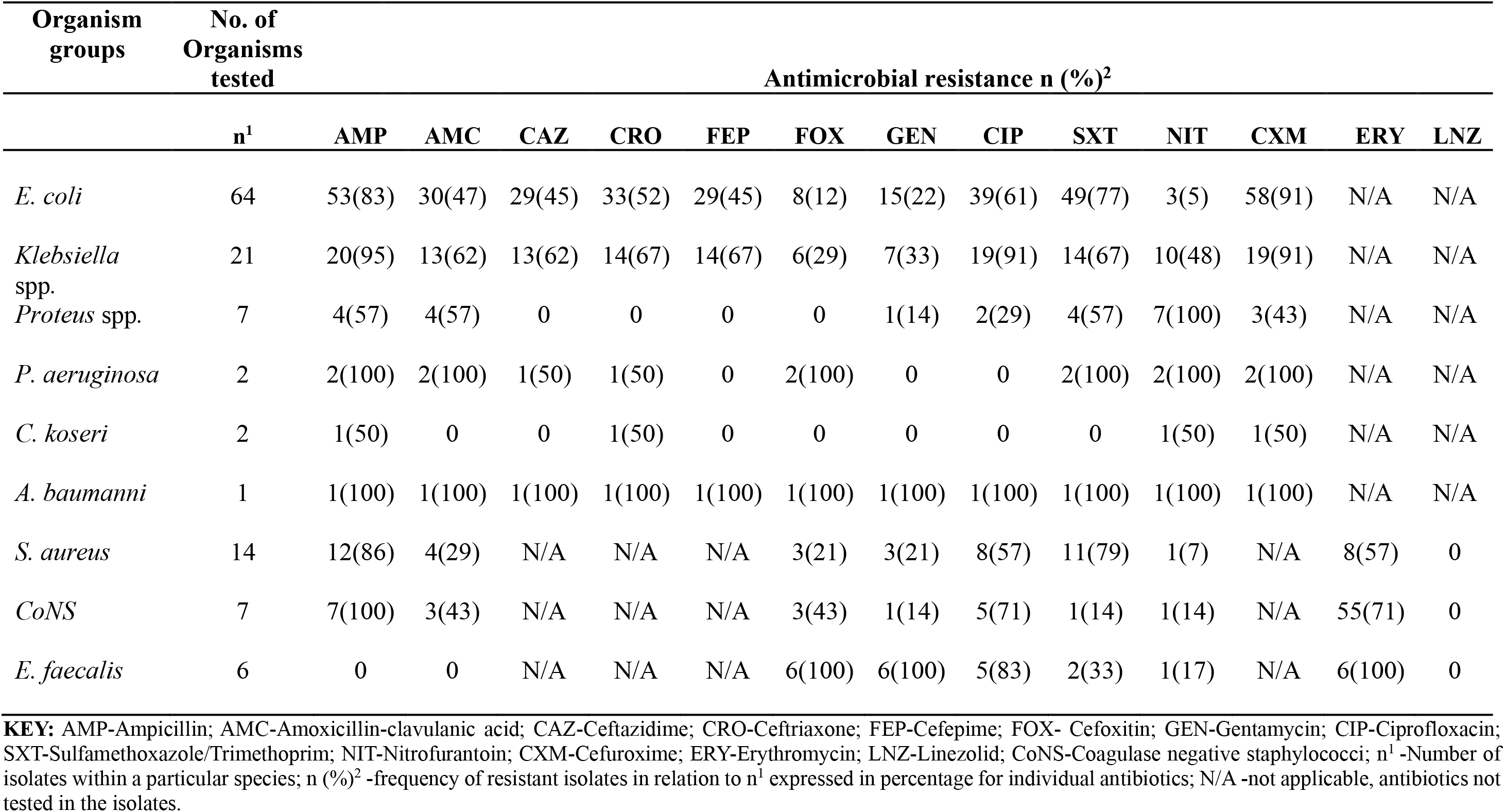
Antimicrobial resistance profiles of isolated UTI pathogens.

### 3.5 Empirical antimicrobial prescribing

There were 15 antibiotics and antibiotic combinations prescribed empirically. Most of the patients 244 (89.0%) received one antibiotic, 28 (10.2%) received 2 antibiotics, while 2 (0.7%) received 3 antibiotics, **Figure 2**. Antimicrobial treatment was prescribed to 49.6% of all patients, with a first line empirical treatment recommended in national guidelines utilised in 29.6% of cases. The most frequently prescribed antibiotics were ciprofloxacin (30.3%), amoxicillin clavulanic acid (17.5%), levofloxacin (15.7%), nitrofurantoin (11.6%), and cefuroxime (10.6%), while the least prescribed were sulfamethoxazole trimethoprim (1.1%), and cefepime at (0.4%). Ceftriaxone/ciprofloxacin and cefixime/azithromycin were the most prescribed combination therapies at 4.7% and 3.2% respectively.

**Figure 2:**
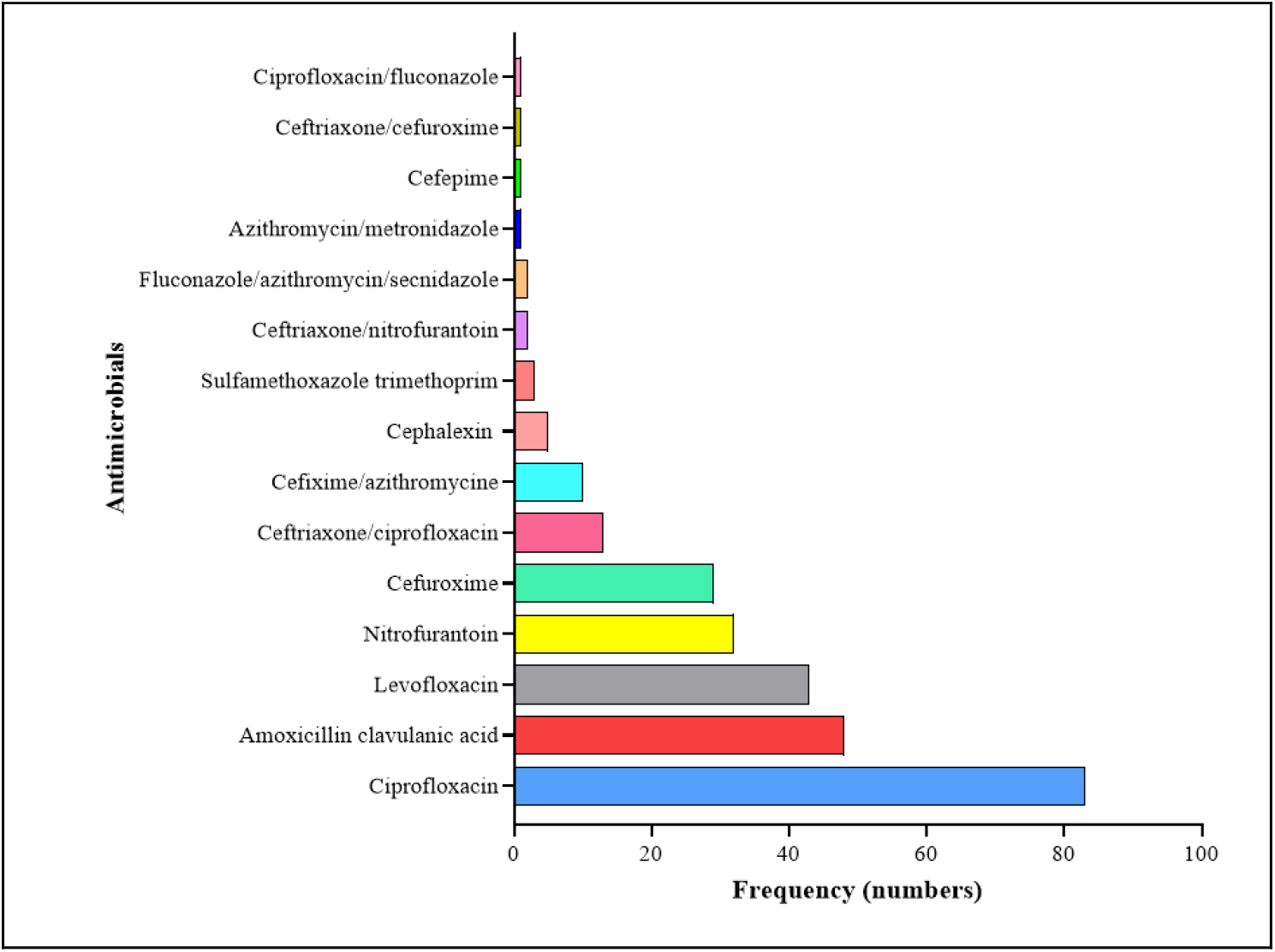
An overview of empirical antimicrobial prescribing at the outpatient departments of MLKH and MCH, Nairobi County, Kenya.

### 3.6. Appropriateness of empirical antibiotic treatment (AEAT)

Of the 95 patients with bacteriological confirmation of UTI, the antimicrobial susceptibility results were compared with the empirical therapy prescribed. The most prescribed antibiotics empirically were found to be inappropriate as follows: Ciprofloxacin was prescribed 27 times but in 11 cases (41%), the isolated organisms were resistant; for amoxicillin clavulanic acid, in 12 (40%) out of the 30 prescriptions, organisms were resistant; for nitrofurantoin, 4 (27%) of the 15 prescriptions proved to be inappropriate, and finally, cefuroxime was prescribed 7 times, but 6 (88%) cases were inappropriate. Overall, most patients 50 (53%) received appropriate empirical therapy, while for 38 (40%), the therapy was found to be inappropriate. The appropriateness of empirical therapy to 7 (7%) patients could not be determined as the antibiotics prescribed (levofloxacin, cefixime/azithromycin) were not in the AST panel, **Figure 3**.

**Figure 3:**
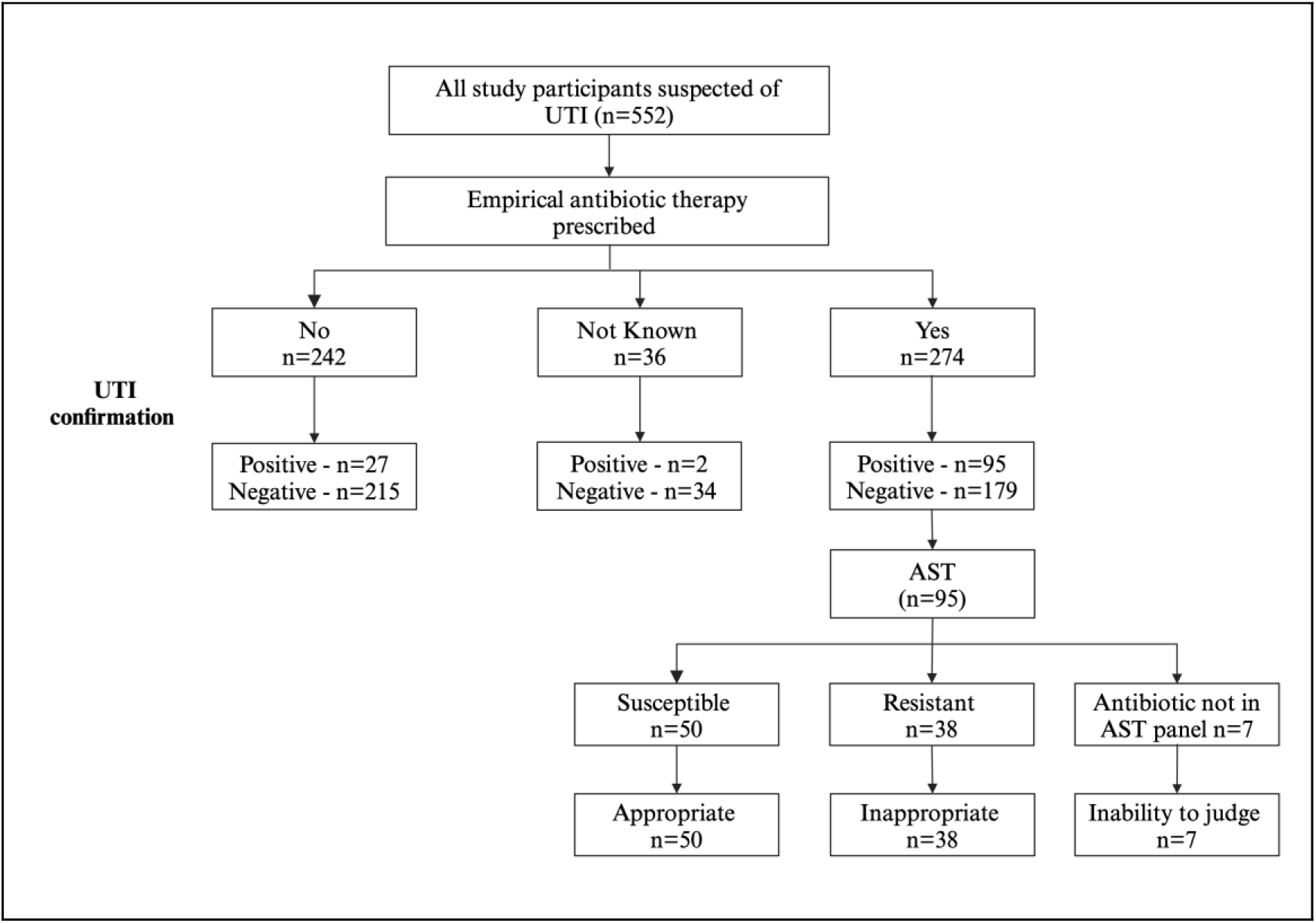
Evaluation of appropriateness of empirical therapy. Appropriateness or inappropriateness was expressed as a percentage based on n=95 patients who had laboratory confirmed UTI and had susceptible or resistant AST results respectively.

## 4. Discussion

This study determined the proportion of microbiologically confirmed UTI cases among 552 symptomatic patients and evaluated the appropriateness of empiric antibiotic therapy prescribed to symptomatic UTI patients. Our findings suggest that in about 40% of cases, empirical antimicrobial prescribing for UTI proves inappropriate in the context of subsequent urine culture and susceptibility results. IEAT can be associated with significant adverse outcomes. While changing to the right antibiotic upon receipt of the culture results is beneficial and necessary for targeted therapy, it may not fully mitigate the disadvantages of not having the correct antibiotic from the onset.^23^ IEAT may promote selection pressures that can result in the growth of resistant bacterial populations, which not only affects the individual patient, but also poses a broader public health threat to everyone. Furthermore, IEAT can result in unnecessary healthcare costs including expenses associated with additional tests and treatment for complications.^24^ These consequences underscore the importance of judicious antibiotic prescribing to optimize patient outcomes and preserve the effectiveness of antibiotics for future generations.

It is challenging to find comparable studies because of the wide variation in the way that IEAT is defined. Nevertheless, a recent study by Maina *et al* investigated the appropriateness of antibiotic use across a range of disease conditions among 1502 patients in Kenyan public hospitals. Among other findings, these results showed that 26% of 94 patients who had UTI and 68% of 135 patients in the surgical unit received empirical treatment that was inappropriate for the pathogens isolated.^25^ Higher rates ranging from 54% to 87% of IEAT in UTI have been reported by other studies.^26–30^ However, while our study defined inappropriate treatment according to the criteria outlined by Davey *et al,*^23^ these studies had a combination of definitions which included: antibiotic prescriptions without bacteriological confirmation, prescription of an antibiotic to which isolated pathogen was resistant, inappropriate antibiotic dosage, lack of sensitivity testing, and therapy not being within the treatment guidelines.

Overall, only 1 in 5 patients suspected of having UTI had bacteriological confirmation by the criteria applied in this study (monoculture growth of 10^4 cfu/ml). However, a considerable proportion of the patients 168 (30%), had taken antibiotics prior to the initial hospital visit. This highlights the challenge of conducting and interpreting microbiology culture results in patients previously exposed to antibiotics, as prior research has demonstrated that antibiotic exposure is a strong predictor of negative culture outcomes.^31^ This further illustrates the difficulty healthcare providers face in deciding on the need for antibiotic prescriptions based solely on clinical symptoms. Evidence on how well symptoms predict the true presence of UTI when compared with urine culture has shown varied results, and is estimated to have an error rate of up to 33%.^31^ In this study, 11% of patients had laboratory confirmation of UTIs, yet they did not receive empirical treatment. These findings are comparable to those reported by Zhue *et al* and Rama *et al*, in which 15.7% and 12.5% patients respectively, did not receive empirical treatment but were confirmed to have UTI by the culture method.^26,29^ While treating only after the microbiological results are obtained ensures that the correct antimicrobial therapy is chosen, the strategy increases the risk of a worse outcome. These findings highlights the need of a near point-of-care test that can detect UTI and provide preliminary antimicrobial susceptibility reports to guide decision-making in UTI management.

There was a wide variation of empirical antimicrobial prescribing practise among prescribers, with differences in preference for certain antimicrobials seen. This was most striking in relation to the prescription of fluoroquinolones (ciprofloxacin, levofloxacin), β-lactam/β-lactamase inhibitor combinations (AMC), and nitrofurantoin. Despite being a second line therapy, more than half 140 (51%) of the patients received fluoroquinolones. This is high considering the already reported high resistance^32^ and adverse ecological effects^33^ associated with this class of antimicrobials. A further 6.3% of the patients received sulfamethoxazole trimethoprim despite this antibiotic not being among the recommended empirical treatments,^16^ and local resistance patterns already exceeding 20%.^32^ One possible explanation to these findings is the absence of sufficient laboratory support, which influences prescription pattern and choice, leading to a predominance of broad-spectrum prescriptions and polypharmacy.^34^ The high-grade resistance exhibited against amoxicillin-clavulanic-acid makes this agent suboptimal for UTI treatment in the absence of laboratory support, notwithstanding that it is recommended in the national guidelines as first-line empirical therapy. This illustrates a clear need for more comprehensive national surveillance and perhaps a review of the guidelines. In contrast, nitrofurantoin was an appropriate agent for both Gram-positive and Gram-negative bacteria, and its empirical use is encouraged in the absence of any contraindication.^35^

The high proportion of resistance amongst UTI pathogens reported in this study and in neighbouring countries,^17,36^ can likely be explained by records of inappropriate antibiotic use which is one of the key drivers of AMR. This could be caused by inadequate microbiology diagnostics, lack of updated antibiotic susceptibility data, and self-treatment using over-the-counter antibiotics, a wide-spread practise in many LMICs.^37^ Some challenges identified in laboratory diagnostics have been the long turnaround time (TAT), high cost of investigation and the lack of trust in and utilization of laboratory results by clinicians.^38^ Performing culture and susceptibility tests may contribute to higher healthcare costs for patients. However, its essential to consider this added expense in light of the potential savings from avoiding inappropriate or unnecessary treatment that are not supported by laboratory data.

This study has some limitations. First, the patients were only recruited from the outpatient departments of two health facilities, so generalisation of findings to other settings, even within Kenya, should be made with caution. Nevertheless, patients were sequentially recruited without stringent selection criteria and the same approach was taken to investigation of every participant which minimised bias and increased the likelihood that the results reflected the general population and routine medical practices. Further, the findings do not give insights into the appropriateness of prescription in private health facilities or in inpatients. However, it is considered satisfactory to provide background information on appropriateness of empirical treatment. Secondly, the population of outpatients who presented with symptoms suggestive of UTI may have had other underlying conditions given that UTI symptoms may over-lap with those of other diseases. However, we assumed that all antibiotics prescribed during initial hospital visit before the AST results (when each patient was recruited into the study and urine collected) were for the UTI episode.

## 5. Conclusion

The study has demonstrated that achieving appropriate empirical antibiotic treatment for UTIs is a difficult task, especially in the era of increased AMR in clinical infections, situations of limited resource, and much habitual over-the counter antibiotic use. At present, optimal empiric therapy is not being achieved. This situation could be improved if capacity for delivering accurate and timely susceptibility results to clinicians to aid their clinical decision making could be achieved. Finally, it is crucial to enhance routine AMR surveillance to support effective antimicrobial stewardship practices in healthcare facilities.

## Data Availability

All data produced in the present work are contained in the manuscript

## Authors contribution

HAO.: conceptualization, laboratory investigations, data curation, formal analysis, and original drafting of the manuscript. RJHH.: Conceptualization, supervision, reviewing and editing. DS.: Conceptualization, supervision, reviewing and editing MK.: Conceptualization, supervision, reviewing and editing KK.: Conceptualization, supervision, reviewing and editing. CWN.: Supervision of laboratory data collection in Kenya. HG.: Laboratory investigations and data curation.

## Acknowledgements

The authors would like to thank all patients for their participation, and the laboratory and clinical teams at MLKH, MDH and KEMRI-CMR for their support during the study period.

## Transparency declarations

None to declare

